# Heterogeneity across race and ethnicity for menopause onset

**DOI:** 10.64898/2026.06.28.26356782

**Authors:** Sarika Pasumarthy, Maggie Hurley, Irene Y. Chen, Nitya Thakkar, Monica Agrawal, Yulin Hswen

**Affiliations:** Electrical Engineering and Computer Science, University of California Berkeley; Epidemiology and Biostatistics, University of California San Francisco; Computational Precision Health, University of California Berkeley and University of California; Berkeley AI Research, University of California Berkeley; Computer Science, Stanford University, Stanford, CA, USA; Duke University Department of Biostatistics and Bioinformatics, Duke University Department of Computer Science; Epidemiology and Biostatistics, University of Maryland School of Public Health; Computer, Mathematics and Natural Sciences, University of Maryland; The Artificial Intelligence Interdisciplinary Institute at Maryland

## Abstract

The heterogeneity onset of menopause varies across racial and ethnic groups, yet this heterogeneity may partially reflect methodological differences rather than true biological differences. Using the All of Us Research Program Controlled Tier dataset (v8), we analyzed age at first menopause diagnosis across three progressively refined cohorts: a full ICD-based cohort (Cohort 1, *n* = 11,306), a survey-linked subcohort adjusted for neighborhood deprivation, smoking, and alcohol use (Cohort 2, *n* = 10,639), and a confirmatory sensitivity cohort applying SNOMED-based surgical exclusions to the same adjusted framework (Cohort 3, *n* = 10,222). Asian & Pacific Islander and Indigenous/Other individuals experienced significantly earlier menopause onset than White individuals across both adjusted cohorts. The Black-White heterogeneity was attenuated after covariate adjustment and did not re-emerge after surgical exclusion. Current smoking was the sole significant behavioral predictor across adjusted models. These findings demonstrate that menopause definition and cohort selection critically shape estimates across race and ethnicity, and that EHR-based ascertainment combined with structured surgical exclusion yields broadly consistent adjusted results.

## Introduction

Menopause marks the permanent cessation of menstruation and the end of a woman’s reproductive capacity.^1^ Clinically, it is recognized after 12 consecutive months without a menstrual period and reflects the depletion of ovarian follicles and a sustained decline in ovarian hormone production, as codified by the Stages of Reproductive Aging Workshop (STRAW+10) framework.^2^ The menopausal transition, often referred to as perimenopause, begins as levels of estrogen and progesterone fluctuate and gradually decline.^3^ For most women, this transition occurs between the ages of 45 and 55, but substantial variation exists in the timing of menopause, and evidence indicates that race and ethnicity play an important role in shaping this trajectory, however research has been scares and less updated in this area with many studies dating back to over 20 decades.

Previous studies have documented racial and ethnic differences in the age of menopausal onset. A 2001 study saw that Africans, African Americans, and Hispanics of Mexican descent have an earlier age at menopause than Caucasian women.^4^ In a 2008 study by Henderson et al., found that compared with non-Latina White women, Latinas experienced significantly earlier menopause, with the association strongest among women born outside the United States.^6^ A current 2023 findings showed that the predicted median age of menopause in Black women was specifically 1.2 years earlier than in White women.^5^

In a 25-year longitudinal, multi-racial/ethnic cohort study, the observed difference in age at menopause between Black and White women was attenuated after adjustment for health behaviors and socioeconomic factors, including education, employment status, cigarette use, and other social determinants.^3^This observation is consistent with a broader body of evidence indicating that earlier menopause is closely linked to health and socioeconomic inequities shaped by intersecting sociocultural, environmental, behavioral, and biological factors.^7,8^Black, Hispanic, and Native women are disproportionately exposed to conditions associated with earlier menopausal onset.^9^These findings underscore that reproductive aging is influenced not only by biological processes but also by social and environmental contexts.

Age at menopause is a clinically significant metric because it is closely linked to long-term health outcomes. Earlier menopause has been associated with an elevated risk of cardiovascular disease (CVD), osteoporosis, and stroke.^10,11^The decline in estrogen, a hormone with protective effects on the vascular endothelium and bone metabolism, can lead to changes in cholesterol levels, blood vessel function, and bone density.^12,13^Consequently, women who experience menopause at younger ages may face a longer period of increased risk for cardiovascular disease and related conditions. This is particularly important for Black and Hispanic women, who have higher rates of metabolic syndrome, a combination of high blood pressure, elevated blood sugar, and unhealthy cholesterol, that further elevates the risk of heart disease, diabetes, and stroke.^7,8^In addition, experiences of racism and related stress, along with neighborhood factors such as limited access to green space, which often reflect patterns of racial and economic segregation, have been associated with earlier menopause and higher cardiometabolic risk.^7^

These findings highlight the role of social and environmental exposures in shaping differences across race and ethnicity in reproductive aging.

Despite recognition of these inequities, racial and ethnic minority women have historically been underrepresented in clinical and epidemiologic studies of menopause.^5^ Such exclusion has limited the ability to detect and fully characterize racial and ethnic differences in menopausal timing and related health outcomes.^14^ Moreover, cohort selection processes may inadvertently exclude individuals who have experienced cumulative social and biological stress, sometimes described as “weathering,” thereby masking the true extent of disparities.^5,8^ Recent work has also highlighted the importance of symptom burden during perimenopause, with significant variation in healthcare-seeking behavior and diagnosis rates across demographic groups.^15^ In this study, we aim to examine heterogeneity in age at first menopause diagnosis by exploring racial and ethnic differences in reproductive aging. Specifically, we assess variation across racial groups (Asian & Pacific Islander, Black, Indigenous/Other, and White). We aim to clarify how menopause onset differs across groups and to contribute to greater understanding of reproductive aging across a diverse population sample.

## Methods

### Study population and data source

We used data from the All of Us Research Program Controlled Tier dataset (v8), a United States-based longitudinal cohort that enrolls diverse participants and links survey responses with electronic health record (EHR) data. The All of Us Research Program is considered a uniquely diverse dataset because it was intentionally designed to move beyond traditional, non-inclusive biomedical research, with over 80% of its participants coming from groups historically underrepresented in medical research making it ideal to study heterogeneity in onset of menopause across racial and ethnicity. The study was restricted to participants with a recorded biological sex of female (`sex_at_birth_concept_id = 45878463`) and available EHR data (`has_ehr_data = 1`), who were between the ages of 18 and 80 at the time of data extraction and had no record of death. The All of Us program was designed to prioritize enrollment of populations historically underrepresented in biomedical research, making it well-suited to the study of racial and ethnic disparities in reproductive aging.^17^

### Menopause identification and cohort definitions

Menopause diagnoses were identified from condition occurrence records using SNOMED standard concepts containing the substring “menopaus,” excluding peri- and postmenopausal terms. Age at first menopause diagnosis was computed as the difference in days between the earliest qualifying condition date and the participant’s date of birth, divided by 365.25. To define a clinically meaningful window, we restricted age at first recorded menopause diagnosis to between 40 and 65 years. This yielded the full ICD cohort of *n* = 11,306 (Cohort 1).

Cohorts 2 and 3 are nested within a survey-linked subcohort restricted to individuals who also appeared in the All of Us `ds_survey table`. Cohort 2 (*n* = 10,639 after complete-case exclusions) retains ICD-based menopause onset as the outcome and adds adjustment for behavioral and socioeconomic covariates derived from survey responses. Cohort 3 (*n* = 10,222) is constructed from the Cohort 2 base by additionally excluding individuals with SNOMED-coded surgical procedures, specifically hysterectomy, bilateral oophorectomy, bilateral salpingo-oophorectomy, or bilateral removal of ovary, identified in either `procedure_occurrence` or `condition_occurrence` records using standard SNOMED concepts (`standard_concept = ‘S’, vocabulary_id = ‘SNOMED’`). A total of 442 individuals (3.9% of the Cohort 2 base) were excluded by this criterion. Cohort 3 is framed as a prespecified confirmatory sensitivity analysis to assess whether the Cohort 2 differences in pattern was robust to the presence of surgically menopausal individuals in the analytic sample. Precise phenotyping of menopause type is critical in EHR-based research, as conflating natural and surgical cases can distort heterogeneity estimates.^18^

### Covariates

Race was self-reported at enrollment and categorized as White (reference), Black, Asian & Pacific Islander (combining Asian and Native Hawaiian or Other Pacific Islander), and Indigenous/Other (combining American Indian or Alaska Native, Middle Eastern or North African, more than one population, and unknown/other). Ethnicity was derived from the All of Us enrollment survey and categorized as Hispanic or Non-Hispanic, with classification based on whether responses contained “Hispanic” but not “Not Hispanic.” Covariates in Cohorts 2 and 3 included smoking status (never [reference], former, current; derived from concept IDs 1585857 and 1585860), alcohol use (moderate [reference], none, heavy; concept IDs 1586198, 1586201, 1586213), and the neighborhood deprivation index from the All of Us zip3-level socioeconomic mapping table (`zip3_ses_map`), treated as a continuous variable using the most recent observation per participant.

### Statistical analysis

Multivariable ordinary least squares (OLS) regression models were fit in Cohorts 2 and 3 with age at first ICD menopause diagnosis as the continuous outcome. White was specified as the reference category for race, consistent with prior literature.^3,5,16^ Adjusted group means were estimated via model prediction, generating predicted values for each race group while holding all other covariates at their reference categories or sample mean for continuous variables. Coefficient comparisons between Cohort 2 and Cohort 3 (Δ*β*) were calculated as the arithmetic difference in point estimates to assess the sensitivity of heterogenous estimates to SNOMED-based surgical exclusion. All analyses were conducted using Python (`pandas, statsmodels, scipy`) within the All of Us Researcher Workbench.

## Results

### Cohort 1: Full ICD cohort (unadjusted)

The full ICD cohort (Cohort 1, *n* = 11,306) had a mean menopause onset age of 52.55 years (median: 52.14, SD: 5.58). White individuals had the highest mean onset age (52.86 years), followed by Black (52.41), Indigenous/Other (51.59), and Asian & Pacific Islander (50.85) individuals (Table 1). The overall mean onset age is consistent with the SWAN median of 52.54 years^3^ and comparable to published multi-ethnic EHR estimates, with variation across groups broadly aligned with prior literature.^4,5^ Despite variation across groups, the overall effect sizes were modest, indicating distributional shifts rather than substantively distinct onset patterns across groups.

**Table 1:** Menopause onset age by race group across all three cohorts.

| Cohort 1 ($n = 11,306$) | | | |
| --- | --- | --- | --- |
| Race Group | n | Mean (SD) | Median [IQR] |
| White | 7,615 | 52.86 (5.63) | 52.43 [48.92–56.76] |
| Black | 1,535 | 52.41 (5.31) | 52.10 [48.88–55.94] |
| Asian & Pacific Islander | 205 | 50.85 (5.10) | 50.87 [47.34–53.50] |
| Indigenous/Other | 1,951 | 51.59 (5.48) | 51.24 [47.77–54.88] |
| Overall | 11,306 | 52.55 (5.58) | 52.14 [48.68–56.29] |
| Cohort 2 ($n = 10,639$) | | | |
| Race Group | n | Mean (SD) | Median [IQR] |
| White | 7,404 | 52.89 (5.64) | 52.47 [48.94–56.81] |
| Black | 1,491 | 52.41 (5.30) | 52.10 [48.88–55.91] |
| Asian & Pacific Islander | 200 | 50.92 (5.09) | 50.91 [47.35–53.51] |
| Indigenous/Other | 1,544 | 51.44 (5.37) | 51.07 [47.75–54.66] |
| Overall | 10,639 | 52.58 (5.57) | 52.16 [48.73–56.33] |
| Cohort 3 ($n = 10,222$) | | | |
| Race Group | n | Mean (SD) | Median [IQR] |
| White | 7,126 | 52.86 (5.62) | 52.45 [48.94–56.75] |
| Black | 1,405 | 52.44 (5.29) | 52.10 [48.90–55.91] |
| Asian & Pacific Islander | 195 | 50.93 (5.07) | 50.91 [47.37–53.53] |
| Indigenous/Other | 1,496 | 51.45 (5.34) | 51.11 [47.82–54.66] |
| Overall | 10,222 | 52.56 (5.56) | 52.15 [48.74–56.28] |

**Figure 1:**
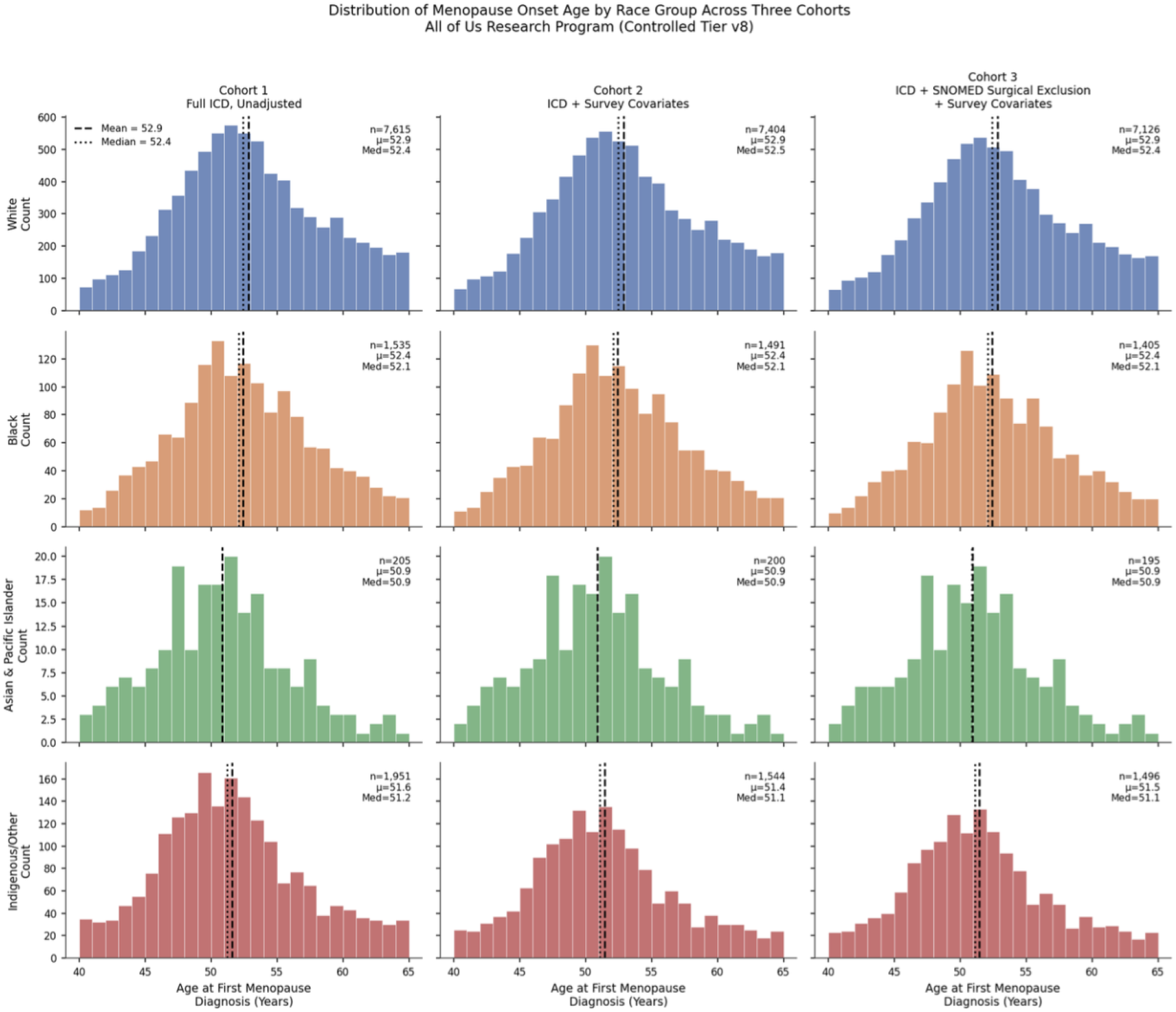
Distribution of menopause onset age by race group across three cohorts, All of Us Research Program (Controlled Tier v8). Each panel shows 1-year bin histograms with mean (dashed line) and median (dotted line) marked. Y-axes are shared within each race row for comparability across cohorts; x-axis is shared across all panels.

### Cohort 2: Survey-linked subcohort, adjusted for SES, smoking, and alcohol

Cohort 2 (*n* = 10,639) was restricted to individuals with both ICD-based menopause diagnoses and completed All of Us lifestyle surveys, retaining those with complete data on smoking status, alcohol use, and neighborhood deprivation index. The mean onset age in this cohort was 52.58 years (median: 52.16), nearly identical to the Cohort 1 estimate, indicating minimal selection bias from the survey linkage requirement. Missing data rates were low across all covariates (smoking: 2.4%, alcohol: 1.3%, deprivation index: 3.1%), and complete-case analysis was applied.

The overall OLS model was statistically significant (*F* = 20.39, *p* = 6.27 × 10^−31^, *R*^2^ = 0.0151, Adj. *R*^2^ = 0.0144, *N* = 10,639). After covariate adjustment, Asian & Pacific Islander individuals experienced menopause 2.06 years earlier than White individuals (*β* = −2.056, 95% CI: −2.838 to −1.273; *p* < 0.001), representing the largest adjusted race heterogeneity in this cohort. Indigenous/Other individuals experienced onset 1.44 years earlier than White individuals (*β* = −1.438, 95% CI: −1.756 to −1.120; *p* < 0.001). The Black-White heterogeneity was attenuated after adjustment and did not reach conventional significance (*β* = −0.307, 95% CI: −0.635 to 0.021; *p* = 0.067), consistent with SWAN findings where the Black-White difference was reduced after controlling for education, employment, and smoking.^3^

Among behavioral predictors, current smoking was significantly associated with earlier menopause (*β* = −1.112 years, 95% CI: −1.491 to −0.734; *p* < 0.001), consistent with established dose-response relationships and aligning closely with SWAN estimates of approximately 1.33 years earlier onset for current smokers.^3^ Former smoking, alcohol use, and the neighborhood deprivation index were not significantly associated with menopause timing in this cohort. Detailed covariate distributions by race group are provided in Table 2.

**Table 2:** Covariate distribution by race group, Cohort 2 (n = 10,639). Values are percentages unless otherwise noted.

| | White<br>($n=7,404$) | Black<br>($n=1,491$) | Asian & PI<br>($n=200$) | Indigenous/Other<br>($n=1,544$) | Overall<br>($n=10,639$) |
| --- | --- | --- | --- | --- | --- |
| Hispanic (%) | 1.5 | 1.1 | 2.0 | 74.2 | 12.0 |
| Smoking: Never (%) | 61.3 | 65.1 | 85.5 | 73.8 | 64.1 |
| Smoking: Former (%) | 31.5 | 15.3 | 12.5 | 17.6 | 26.8 |
| Smoking: Current (%) | 7.2 | 19.7 | 2.0 | 8.7 | 9.1 |
| Alcohol: None (%) | 2.5 | 14.2 | 22.5 | 21.2 | 7.2 |
| Alcohol: Moderate (%) | 95.7 | 82.8 | 77.0 | 76.9 | 90.8 |
| Alcohol: Heavy (%) | 1.9 | 3.0 | 0.5 | 1.9 | 2.0 |
| Deprivation Index, Mean (SD) | 0.299 (0.054) | 0.347 (0.063) | 0.299 (0.069) | 0.325 (0.066) | 0.310 (0.060) |
| Onset Age, Mean (SD) | 52.89 (5.64) | 52.41 (5.30) | 50.92 (5.09) | 51.44 (5.37) | 52.58 (5.57) |
| Onset Age, Median [IQR] | 52.47<br>[48.94–56.81] | 52.10<br>[48.88–55.91] | 50.91<br>[47.35–53.51] | 51.07 [47.75–54.66] | 52.16<br>[48.73–56.33] |

**Figure 2:**
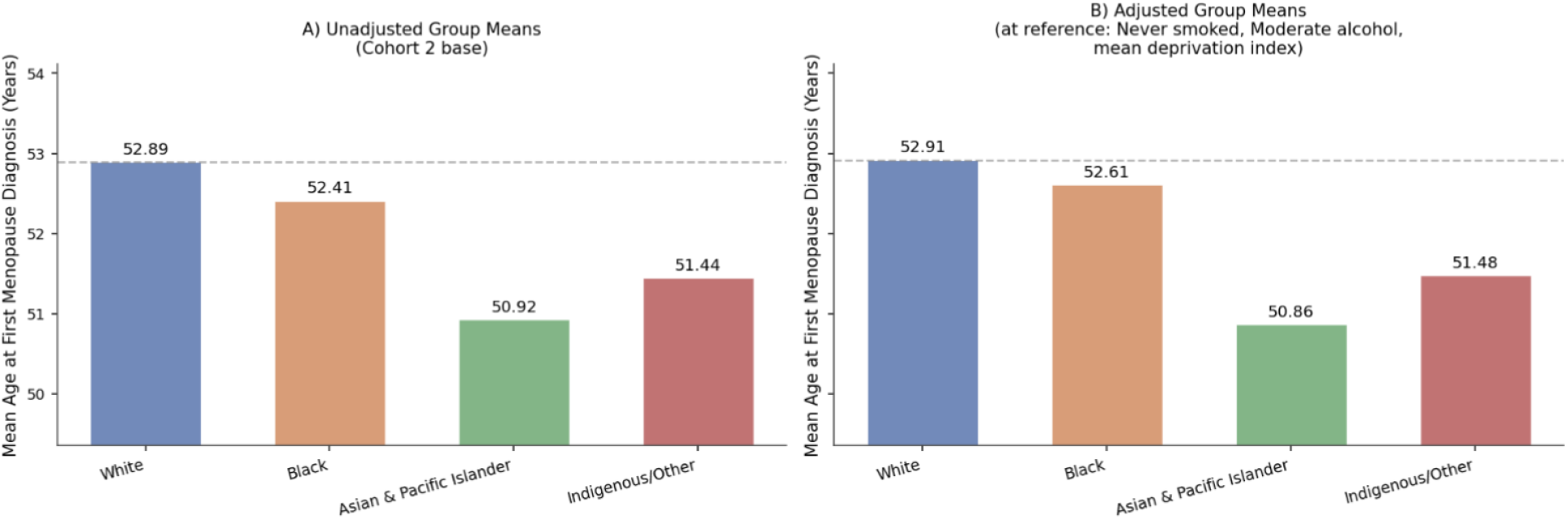
Unadjusted versus adjusted mean menopause onset age by race group (Cohort 2, ICD-based, n = 10,639). Panel A shows unadjusted group means; Panel B shows model-predicted means standardized to the reference covariate profile (never smoked, moderate alcohol, sample mean deprivation index). Near-convergence of White and Black adjusted means illustrates attenuation of the Black-White heterogeneity after covariate adjustment.

**Figure 3:**
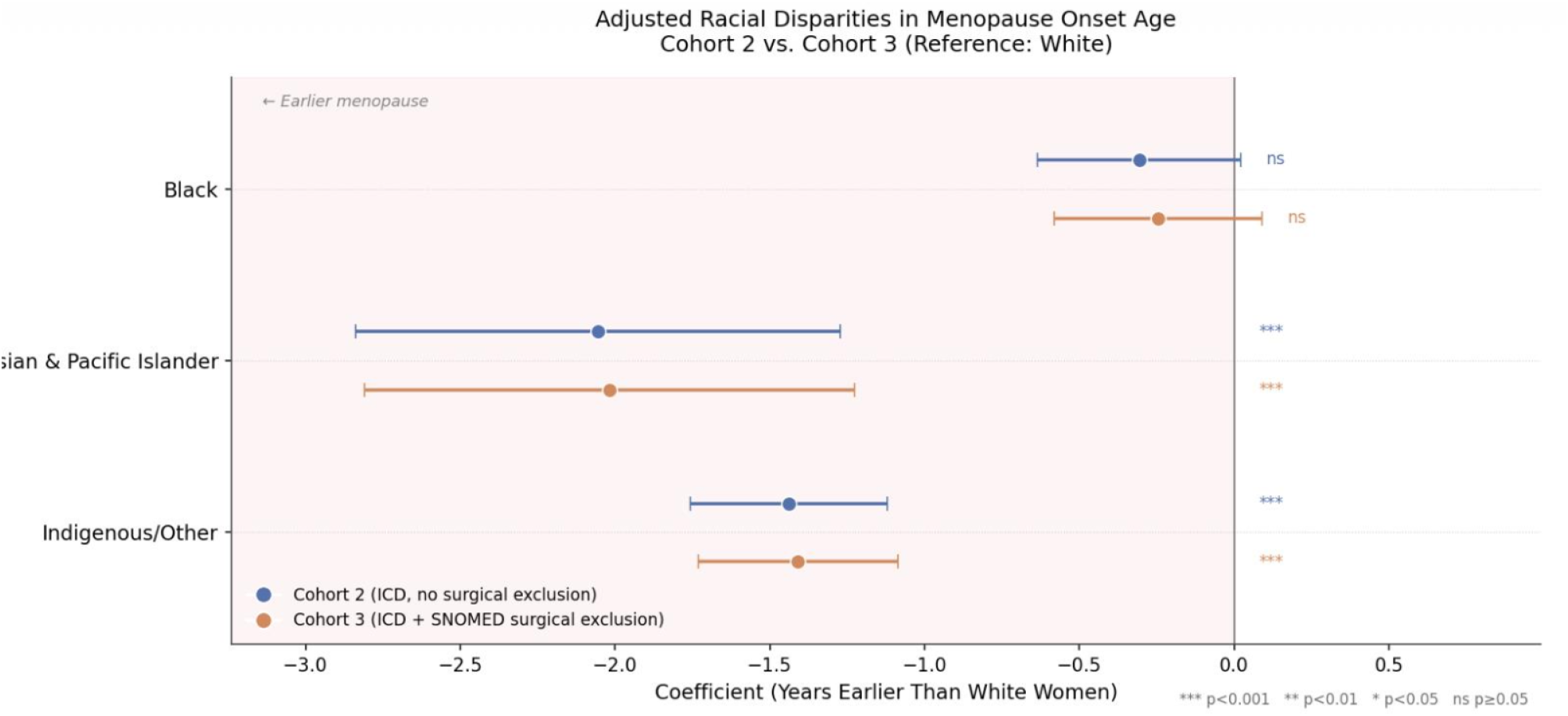
Forest plot of adjusted race group coefficients with 95% confidence intervals for Cohort 2 (blue) and Cohort 3 (orange), reference group: White. Point estimates represent the adjusted difference in menopause onset age (years) relative to White women. The dashed vertical line at zero indicates no difference. Asterisks denote significance: *** *p* < 0.001; ns, not significant.

### Cohort 3: ICD menopause with SNOMED-based surgical exclusion (sensitivity analysis)

To evaluate the robustness of Cohort 2 findings to the potential confounding influence of surgical menopause, we constructed Cohort 3 as a prespecified confirmatory sensitivity analysis. Starting from the Cohort 2 base, we excluded individuals with SNOMED-coded hysterectomy, bilateral oophorectomy, or bilateral salpingo-oophorectomy identified in either `procedure_occurrence or condition_occurrence` records. This resulted in the exclusion of 442 individuals (3.9% of the Cohort 2 base), yielding a Cohort 3 analytic sample of *n* = 10,222 after complete-case exclusions.

The mean onset age in Cohort 3 was 52.56 years (median: 52.15, SD: 5.56), virtually identical to Cohort 2 (52.58 years), indicating that the small proportion of surgically menopausal individuals identified via SNOMED codes did not materially shift the overall onset distribution. The overall model fit was similarly strong (*F* = 19.07, *p* = 9.77 × 10^−29^, *R*^2^ = 0.0147, Adj. *R*^2^ = 0.0139, *N* = 10,222).

Adjusted race group coefficients in Cohort 3 were strikingly consistent with Cohort 2 (Table 3). Asian & Pacific Islander individuals experienced onset 2.02 years earlier than White individuals (*β* = −2.018, 95% CI: −2.809 to −1.227; *p* < 0.001). Indigenous/Other individuals experienced onset 1.41 years earlier (*β* = −1.409, 95% CI: −1.731 to −1.086; *p* < 0.001). The Black-White coefficient remained attenuated and non-significant (*β* = −0.246, 95% CI: −0.582 to 0.090; *p* = 0.152). The change in the Black-White coefficient from Cohort 2 to Cohort 3 (Δ*β* = +0.061 years) was negligible, indicating that the 442 excluded individuals with SNOMED surgical codes did not substantively alter the racial heterogeneous pattern. Current smoking replicated as the sole significant behavioral predictor (*β* = −1.175, 95% CI: −1.559 to −0.790; *p* < 0.001), essentially unchanged from Cohort 2.

**Table 3:**
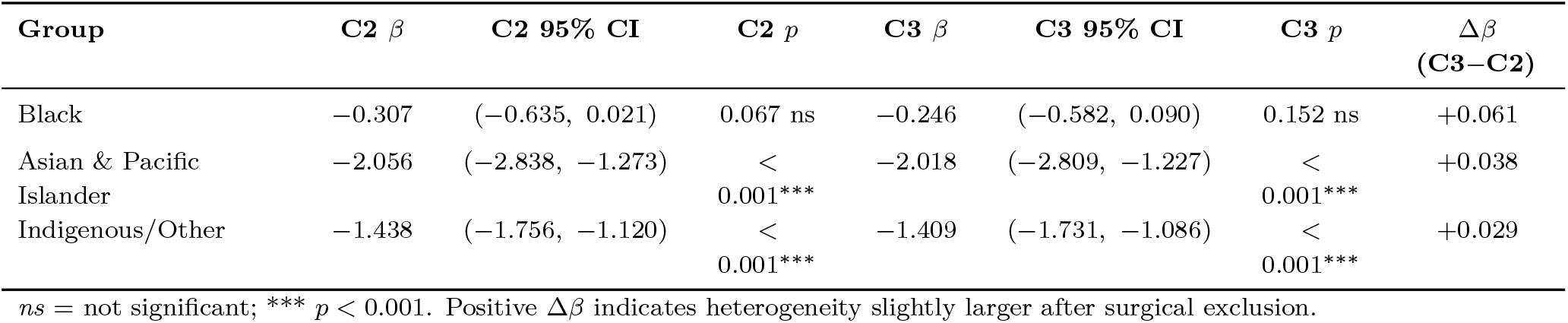
Adjusted race group coefficients from OLS models, Cohort 2 vs. Cohort 3 (reference: White).

**Figure 4:**
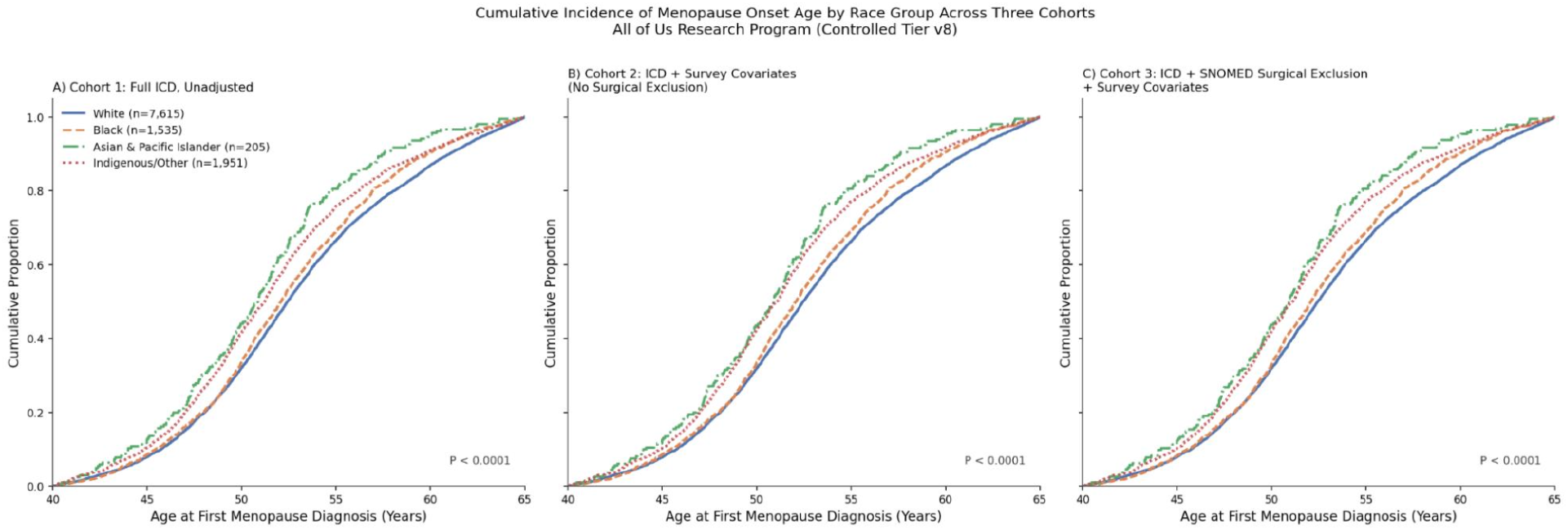
Cumulative incidence of menopause onset age by race group across three cohorts, All of Us Research Program (Controlled Tier v8). Kruskal-Wallis p < 0.0001 for all three cohorts. Curves are directly comparable as all three cohorts use the same ICD-based outcome (`first_meno_icd_age`).

**Figure 5:**
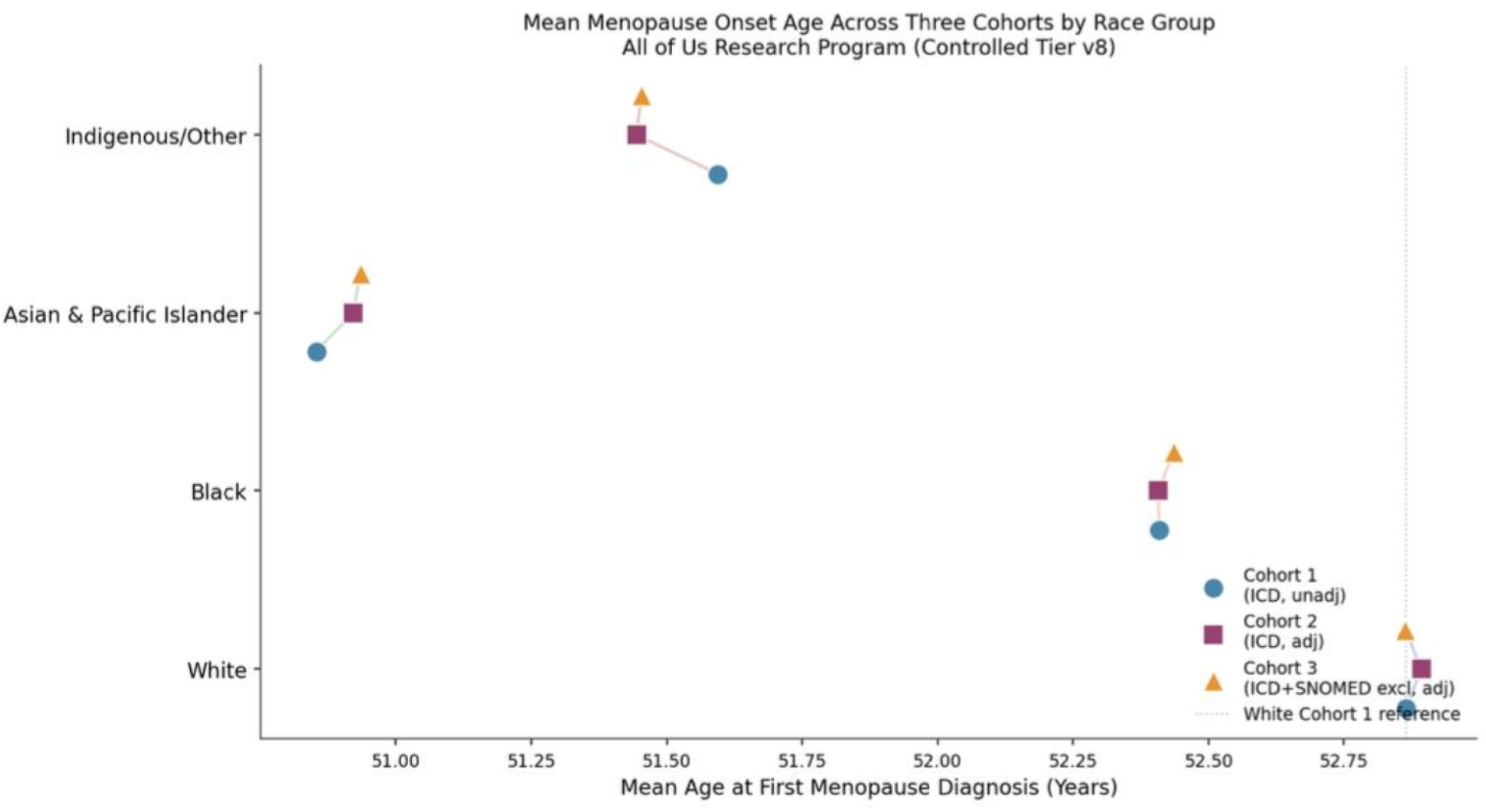
Mean menopause onset age by race group across three cohorts. Each race group is shown on the y-axis; cohort membership is denoted by marker shape and color. Connecting lines within each race group illustrate the stability of group-level estimates across cohort refinement steps.

## Discussion

This study demonstrates that heterogeneity across race and ethnicity in menopause timing are robust and persist after adjustment for key behavioral and socioeconomic factors, though their magnitude depends on menopause ascertainment strategy and cohort selection. By examining the same population through three progressively refined cohort definitions, full ICD ascertainment, survey-linked adjustment, and SNOMED-based surgical exclusion, we show that different estimates across groups are meaningfully stable across analytic refinement steps, with the largest difference of menopause onset concentrated in Asian & Pacific Islander and Indigenous/Other groups relative to White women.

Asian & Pacific Islander individuals showed the largest and most consistent adjusted differences in menopause onset across both Cohort 2 (*β* = −2.06 years, *p* < 0.001) and Cohort 3 (*β* = −2.02 years, *p* < 0.001). This magnitude is larger than estimates from some prior studies, likely reflecting heterogeneity within this aggregated group of Asians. Henderson et al.^6^ found that Japanese Americans had later menopause than non-Latina White women, underscoring the need for disaggregated analyses within this broad grouping. Recent evidence further demonstrates substantial heterogeneity within the aggregated “Asian” category, with menopause timing and related health outcomes varying significantly across subgroups and being strongly influenced by socioeconomic status, reinforcing the importance of disaggregating this population in epidemiologic analyses.^22^ The stability of the Asian & Pacific Islander heterogeneity across both adjusted cohorts, with Δ*β* of only 0.038 years between Cohort 2 and Cohort 3, indicates that this finding is not an artifact of differential surgical menopause rates. Indigenous/Other individuals similarly showed a robust and consistent adjusted heterogeneity of approximately 1.4 years earlier onset across both cohorts (Cohort 2: *β* = −1.438, *p* < 0.001; Cohort 3: *β* = −1.409, *p* < 0.001). This group is heterogeneous and includes a substantial proportion of individuals identifying as Hispanic (74.2% in Cohort 2), which likely contributes to the earlier onset observed in this category.

The Black-White heterogeneity followed a different trajectory across the three cohorts. In Cohort 1, Black individuals had a mean onset age 0.45 years earlier than White individuals (52.41 vs. 52.86 years). After covariate adjustment in Cohort 2, this difference was attenuated and did not reach conventional significance (*β* = −0.307, *p* = 0.067), replicating SWAN’s finding that racial differences in menopause timing were reduced after controlling for education, employment, and smoking.^3^ Critically, in Cohort 3, after excluding 442 individuals with SNOMED-coded surgical procedures, the Black–White estimate moved only marginally toward the null (*β* = −0.246, *p* = 0.152; Δ*β* = +0.061 years), remaining non-significant. This finding diverges from the pattern documented by Reeves et al.^5^, who demonstrated that surgical exclusion caused the Black-White heterogeneity to re-emerge in SWAN. The most likely explanation is that the SNOMED-based surgical exclusion applied here, while methodologically rigorous within the EHR context, captured only 3.9% of the Cohort 2 base (*n* = 442), a substantially smaller proportion than the 30.9% surgical menopause rate documented in SWAN for Black women.^5^ It is plausible that a meaningful number of surgically menopausal individuals remain in our Cohort 3 sample due to incomplete EHR capture of surgical procedures, particularly for procedures performed outside of health systems contributing data to All of Us. This interpretation is consistent with Babbar’s commentary, which underscores the difficulty of distinguishing natural from surgical menopause in structured health data, and with Alba et al., who demonstrated that structured diagnostic codes alone are insufficient to accurately classify menopause type in administrative data.^18,19^

Current smoking was the only behavioral covariate reaching significance across both adjusted cohorts (Cohort 2: −1.11 years, *p* < 0.001; Cohort 3: −1.17 years, *p* < 0.001), replicating well-established findings from SWAN^3^ and REGARDS.^16^ The consistency of this estimate across cohorts and its alignment with published dose-response relationships provide internal validation for the analytic approach. Former smoking, alcohol use, and the neighborhood deprivation index were not significantly associated with menopause timing in either adjusted cohort.

In a recent study, Zhou et al. developed a machine learning model to predict early natural menopause in women in China using 20 predictive factors, achieving an AUC of 0.731.^20^ However, the model was developed exclusively in a Chinese population and was not externally validated across other racial or ethnic groups. Our findings suggest that menopause timing varies meaningfully across populations and is sensitive to cohort definition, indicating that predictive models may not generalize well without accounting for racial and ethnic heterogeneity. Separately, a large UK Biobank analysis found that each additional year in age at menopause was associated with a 1.4% increase in all-cause mortality risk, reinforcing the long-term clinical significance of variation in menopause timing and the importance of identifying groups at elevated risk for earlier onset.^21^ Cunningham et al. further documented substantial variation in perimenopause symptom severity and healthcare-seeking behavior across demographic groups in the US, consistent with the differential ascertainment patterns that motivate our multi-cohort design.^15^

Several limitations warrant acknowledgment. Age at first ICD code is a retrospective proxy for menopause onset and may reflect documentation timing rather than true biological onset; symptoms may precede diagnosis by months or years. The SNOMED-based surgical exclusion captures only procedures recorded in the EHR data contributed to All of Us, and may not fully identify individuals who underwent hysterectomy or oophorectomy in health systems not represented in the dataset. The neighborhood deprivation index is an ecological measure at the 3-digit ZIP code level that does not capture individual-level socioeconomic status, and key potential confounders including parity, body mass index, and cumulative stress exposures were unavailable for adjustment. The All of Us cohort, while large and diverse, is not a probability sample and may not be representative of the general U.S. population. Finally, the Asian & Pacific Islander and Indigenous/Other categories aggregate heterogeneous subgroups whose disaggregation may reveal additional variation not detectable at current sample sizes.

In summary, racial and ethnic differencers in menopause timing are robust and clinically meaningful, particularly for Asian & Pacific Islander and Indigenous/Other individuals. The Black–White heterogeneity was attenuated after covariate adjustment and remained non-significant after SNOMED-based surgical exclusion, a finding that may reflect incomplete EHR capture of surgical procedures rather than the absence of a true difference. These findings underscore the importance of precise menopause phenotyping, comprehensive surgical case identification, and careful consideration of cohort selection in epidemiologic research on reproductive aging.

## Ethics

The All of Us Research Program is overseen by an institutional review board, and all participants provided informed consent. This analysis was conducted under the program’s data use agreement and approved protocols.

## Data Availability

The data that support the findings of this study are available from the All of Us Research Program Controlled Tier dataset (v8), but restrictions apply to the availability of these data, which were used under the program’s data use agreement for the current study, and so are not publicly available. Data are available from the All of Us Research Program upon reasonable request and with completion of the program’s registration and data access requirements (https://www.researchallofus.org).

